# Joint Association of Albuminuria and Left Ventricular Hypertrophy with Incident Heart Failure in High-Risk Adults with Hypertension: a SPRINT substudy

**DOI:** 10.1101/2023.07.06.23292329

**Authors:** Muhammad Imtiaz Ahmad, Richard Kazibwe, Mai Z. Soliman, Sanjay Singh, Lin Y. Chen, Elsayed Z. Soliman

## Abstract

**Background:** Albuminuria and left ventricular hypertrophy (LVH) are independent predictors of heart failure (HF), however their combined effect on risk of HF has not been explored previously.

**Objectives:** To examine the joint associations of albuminuria and electrocardiographic (ECG) LVH with incident acute decompensated HF (ADHF), and whether albuminuria/LVH combinations modified the effects of blood pressure control strategy in reducing the risk of ADHF.

**Methods:** 8,511 participants from the SPRINT (Systolic Blood Pressure Intervention Trial) were included. ECG-LVH was present if any of the following criteria: Cornell voltage, Cornell voltage product, or Sokolow Lyon were present. Albuminuria was defined as urine albumin-creatinine ratio (UACR) ≥30 mg/g. ADHF was defined as hospitalization or emergency visit for ADHF. Cox proportional hazard models were used to examine the association of neither LVH, nor albuminuria (reference), either LVH or albuminuria, and both (LVH + albuminuria) with incident ADHF.

**Results:** Over a median follow-up of 3.2 years, 182 cases of ADHF occurred. In adjusted models, concomitant albuminuria and LVH were associated with higher risk of ADHF than either albuminuria or LVH in isolation (HR (95% CI): 4.95 (3.22-7.62), 2.04 (1.39-3.00), and 1.47 (0.93-2.32), respectively (additive interaction *p*=0.01). The effect of intensive blood pressure in decreasing ADHF attenuated among participants with co-existing albuminuria and LVH without any interaction between treatment group assignment and albuminuria/LVH categories (interaction p-value= 0.26).

**Conclusions:** Albuminuria and LVH are additive predictors of ADHF. The effect of intensive blood pressure control in decreasing ADHF risk did not vary significantly across albuminuria/LVH combinations.

URL: https://www.clinicaltrials.gov; Unique identifier: NCT01206062

## Introduction

Left ventricular hypertrophy (LVH) and albuminuria represent interrelated complications of uncontrolled hypertension^1^. Additionally, a complex bidirectional relationship exists between LVH and albuminuria, with each representing a risk factor for the other and vice versa.^2-4^ LVH underlies maladaptive cardiac remodeling due to chronic pressure overload, chronic myocardial injury, inflammation, and fibrosis and has strong associations with incident heart failure (HF) and mortality^5^. Albuminuria is a marker of inflammation and endothelial dysfunction and has been strongly associated with incident HF in multiple independent cohort studies^6,7^. Regression of albuminuria with newer therapies has been associated with clinical benefits such as a reduction in HF hospitalization and mortality^6,8^. Similarly, regression of LVH with antihypertensive therapies reduced stroke risk, myocardial infarction (MI), HF, and mortality^9,10^. Given the significant prognostic nature of these markers, an examination of the joint association of LVH and albuminuria with incident HF may identify very high-risk populations. Further, it is also unclear if intensive blood pressure control would modify the joint association of LVH and albuminuria with incident HF. Therefore, we proposed to examine the association and interaction of albuminuria and electrocardiographic LVH (ECG-LVH) with incident acute decompensated HF (ADHF) using data from the Systolic Blood Pressure Intervention Trial (SPRINT) trial.

## Methods

### Design and Sample

The rationale and design of the SPRINT trial have been previously reported in detail^11^ and a copy of protocol is available in **Supplement 2**. The CONSORT diagram for the study is shown in **Supplemental Figure 1**. Briefly, SPRINT was a randomized, controlled, open-label trial in which 9361 participants ≥50 years of age with systolic blood pressure (SBP) ≥130-180 mmHg and at high risk for or with CVD were randomized to achieve either an SBP target of <140 mmHg (standard treatment group) or <120 mmHg (intensive treatment group). SPRINT aimed to test whether intensive SBP control to <120 mmHg results in the reduction of a composite of CVD events. High CVD risk was defined as ≥1 of the following: clinical or subclinical CVD; chronic kidney disease (CKD); 10-year risk of CVD ≥15% by Framingham risk score; or age ≥75 years. Clinical CVD was defined as the following: 1) prior myocardial infarction (MI), coronary artery bypass grafting, carotid endarterectomy, or carotid stenting; peripheral arterial disease with revascularization; 2) acute coronary syndrome with or without resting ECG change, ECG changes on an exercise test or positive cardiac imaging study; 3) at least a 50% diameter stenosis of a coronary, carotid, or lower extremity artery; or abdominal aortic aneurysm ≥5cm with or without repair. Subclinical CVD was defined as the following: 1) coronary artery calcium (CAC) score ≥400 Agatston units; b) ankle-brachial index (ABI) ≤0.90; 2) LVH by ECG, echocardiogram report, or other cardiac imaging procedure. CKD was defined as estimated glomerular filtration rate (eGFR) <60ml/min/1.73m^2^ using the four-variable Modification of Diet in Renal Disease (MDRD) equation^12^. Participants with a known LV ejection fraction of <35%, any symptomatic HF within 6 months, diabetes mellitus, prior transient ischemic attack or stroke, polycystic kidney disease, dementia, non-adherence to medication, and eGFR <20 mL/min/1.73 m^2^ were excluded. The SPRINT study was approved by the Institutional Review Board at each participating study site, and all participants provided written informed consent. The initial trial was registered at ClinicalTrials.gov (NCT01206062). For the purpose of this analysis, we included 8,511 SPRINT participants with a baseline standard 12-lead ECG and baseline measures of urine albumin creatinine ratio (UACR). We excluded participants with missing ECG or uninterpretable baseline ECG or missing UACR values or a history of atrial fibrillation (AF). A limited de-identified dataset was obtained from the National Heart, Lung, and Blood Institute’s Biologic Specimen and Data Repository Information Coordinating Center after the study was approved by the institutional review board at the Medical College of Wisconsin.

### Electrocardiographic Data

LVH was detected using standard 12-lead ECGs obtained at baseline. Digital ECGs were recorded using a GE MAC 1200 electrocardiograph at 10 mm/mV calibration and a speed of 25 mm/s. ECGs were read centrally at the Epidemiological Cardiology Research Center, Wake Forest School of Medicine, Winston Salem, North Carolina. All ECG tracings were inspected visually for technical errors and inadequate quality before being automatically processed using GE 12-SL Marquette version 2001. LVH was present if met any of the following criteria; Cornell voltage criteria (RaVL amplitude + SV_3_ amplitude) with sex-specific thresholds of ≥2,200 μV in women and ≥2,800 μV in men, or Sokolow-Lyon (SV_1_ amplitude + RV5/V_6_ amplitude) LVH criteria or Cornell voltage product ([RaVL amplitude+SV_3_ amplitude] ×QRS duration)^13^.

### Albuminuria

Albuminuria was estimated by a random urine albumin-creatinine ratio (UACR). Serum creatinine and urine creatinine were measured using an enzymatic method (Roche, Indianapolis, IN) at the SPRINT central laboratory at the University of Minnesota^14^. Albuminuria was defined as UACR ≥30 mg/g.

### Heart Failure Events

Data about potential outcomes were assessed every 3 months in both arms using a standardized protocol with centralized monitoring by the coordinating center. To minimize any ascertainment bias, a structured interview was used to obtain self-reported CVD outcomes^11^. All ADHF events were new or incident HF events and included both HF with reduced ejection fraction (HFrEF) and HF with preserved ejection fraction (HFpEF). ADHF events were adjudicated by a Morbidity and Mortality committee using the standardized Atherosclerosis Risk in Communities (ARIC) study adjudication system^15^. Adjudicators were blinded to treatment assignment. ADHF was defined as a hospitalization or emergency (ED) visit for a clinical syndrome with multiple symptoms and signs consistent with cardiac decompensation and inadequate cardiac pump function requiring treatment with diuretics or inotropic agents. Symptoms supporting the diagnosis of ADHF included evidence of new or increasing shortness of breath, peripheral edema, orthopnea, or paroxysmal nocturnal dyspnea. Positive signs supporting the diagnosis of ADHF included hypoxia, pulmonary rales on clinical examination, pulmonary vascular congestion on chest X-ray, the elevation of B-type natriuretic peptide (BNP) or pro-N-terminal BNP (NTpro-BNP) above the diagnostic threshold, reduced left ventricular ejection fraction or diastolic dysfunction, new or increased treatment with intravenous loop diuretic or inotrope for ADHF, documented response to therapy or evidence in treating physician’s notes that the primary reason for the hospitalization or ED visit was ADHF. Chronic stable HF, right-sided HF, low EF without symptoms of HF, or HF related to volume overload due to inadequate dialysis with end-stage renal disease and new outpatient HF were not included as ADHF endpoints.

### Statistical analyses

Four categories were created as follows: neither LVH, nor albuminuria, either LVH or albuminuria, and both (LVH + albuminuria). The baseline characteristics were compared across LVH/albuminuria categories using analysis of variance for continuous variables, and chi-square test for categorical variables. Continuous variables were reported as mean and standard deviation, median (25%, 75% percentile) for skewed variables, and n (%) for categorical variables.

Incidence rates for ADHF were calculated for each group and Kaplan-Meier plot and the log-rank test were used to compare ADHF-free survival across these groups. We evaluated associations of LVH/albuminuria categories with risk of the ADHF using multivariable Cox proportional hazards models. There was no evidence that the proportional hazards assumptions were violated. Model 1 adjusted for demographics (age, sex, race, randomization site), and model 2 further adjusted for treatment assignment, body mass index, smoking status, prevalent CVD, SBP, eGFR, total cholesterol, statin use, and the number of antihypertensive agents. We tested the additive interaction between LVH and albuminuria and measured additivity by relative excess risk due to interaction (RERI), the proportion of disease attributable to interaction (AP), and synergy index (S)^16^. We also tested the multiplicative interaction between LVH and albuminuria for ADHF risk using the likelihood ratio test in a model that included main effects.

ADHF event rates were compared between the intensive SBP and standard SBP arms among participants in each albuminuria/LVH category. The heterogeneity of treatment effect across albuminuria/LVH categories was tested using a likelihood ratio test for multiplicative interaction terms (treatment assignment by albuminuria/LVH category) in models that included main effects.

As the risk of HF exists even at low-grade albuminuria as well^7^, we used a threshold of UACR ≥10 mg/g and then examined the association of newly created albuminuria/LVH categories with ADHF adjusting for covariates as mentioned above. We also tested multiplicative and additive interaction between albuminuria (UACR ≥10 mg/g) and LVH as described in the main analysis. Additionally, we reported incidence rates of ADHF across UACR categories (<10 mg/g, 10-29.99 mg/g, ≥30 mg/g-299.99 mg/g, and ≥300 mg/g) stratified by LVH status. Using Cox proportional hazard analysis, we computed HR and 95% of association of these UACR categories with incident ADHF stratified by LVH status.

All data analyses were performed in SAS version 9.4 (SAS Institute Inc, Cary, NC), and two-sided α=0.05 was used for hypothesis testing.

## Results

Of 9,361 SPRINT participants, 8,511 with complete ECG and UACR data and free of prevalent AF were included in this analysis. The baseline characteristics of the participants stratified by albuminuria/LVH categories are shown in **Table 1**. In addition to predominant demographic factors such as older age, men, and Black individuals, clinical/subclinical CVD and CVD risk factors were also prevalent among participants with concomitant albuminuria and LVH **(Table 1).**

**Table 1:**
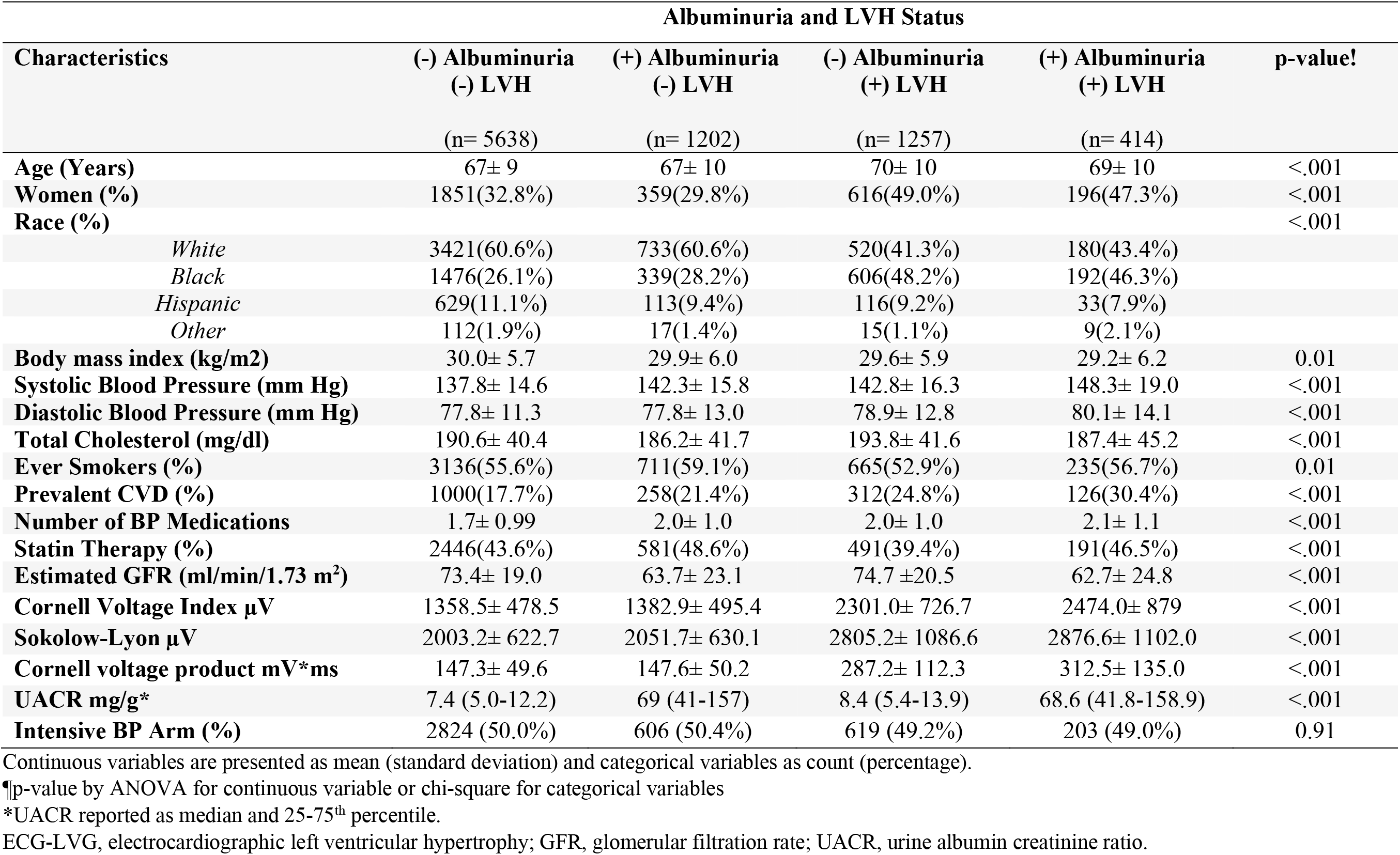
Baseline Characteristics of Study Participants by Albuminuria and LVH Status.

| Characteristics | Albuminuria and LVH Status |  |  |  | p-value! |
| --- | --- | --- | --- | --- | --- |
|  | (-) Albuminuria<br>(-) LVH | (+) Albuminuria<br>(-) LVH | (-) Albuminuria<br>(+) LVH | (+) Albuminuria<br>(+) LVH |  |
|  | (n= 5638) | (n= 1202) | (n= 1257) | (n= 414) |  |
| **Age (Years)** | 67± 9 | 67± 10 | 70± 10 | 69± 10 | <.001 |
| **Women (%)** | 1851(32.8%) | 359(29.8%) | 616(49.0%) | 196(47.3%) | <.001 |
| **Race (%)** |  |  |  |  | <.001 |
| *White* | 3421(60.6%) | 733(60.6%) | 520(41.3%) | 180(43.4%) |  |
| *Black* | 1476(26.1%) | 339(28.2%) | 606(48.2%) | 192(46.3%) |  |
| *Hispanic* | 629(11.1%) | 113(9.4%) | 116(9.2%) | 33(7.9%) |  |
| *Other* | 112(1.9%) | 17(1.4%) | 15(1.1%) | 9(2.1%) |  |
| **Body mass index (kg/m<sup>2</sup>)** | 30.0± 5.7 | 29.9± 6.0 | 29.6± 5.9 | 29.2± 6.2 | 0.01 |
| **Systolic Blood Pressure (mm Hg)** | 137.8± 14.6 | 142.3± 15.8 | 142.8± 16.3 | 148.3± 19.0 | <.001 |
| **Diastolic Blood Pressure (mm Hg)** | 77.8± 11.3 | 77.8± 13.0 | 78.9± 12.8 | 80.1± 14.1 | <.001 |
| **Total Cholesterol (mg/dl)** | 190.6± 40.4 | 186.2± 41.7 | 193.8± 41.6 | 187.4± 45.2 | <.001 |
| **Ever Smokers (%)** | 3136(55.6%) | 711(59.1%) | 665(52.9%) | 235(56.7%) | 0.01 |
| **Prevalent CVD (%)** | 1000(17.7%) | 258(21.4%) | 312(24.8%) | 126(30.4%) | <.001 |
| **Number of BP Medications** | 1.7± 0.99 | 2.0± 1.0 | 2.0± 1.0 | 2.1± 1.1 | <.001 |
| **Statin Therapy (%)** | 2446(43.6%) | 581(48.6%) | 491(39.4%) | 191(46.5%) | <.001 |
| **Estimated GFR (ml/min/1.73 m<sup>2</sup>)** | 73.4± 19.0 | 63.7± 23.1 | 74.7 ±20.5 | 62.7± 24.8 | <.001 |
| **Cornell Voltage Index μV** | 1358.5± 478.5 | 1382.9± 495.4 | 2301.0± 726.7 | 2474.0± 879 | <.001 |
| **Sokolow-Lyon μV** | 2003.2± 622.7 | 2051.7± 630.1 | 2805.2± 1086.6 | 2876.6± 1102.0 | <.001 |
| **Cornell voltage product mV\*ms** | 147.3± 49.6 | 147.6± 50.2 | 287.2± 112.3 | 312.5± 135.0 | <.001 |
| **UACR mg/g\*** | 7.4 (5.0-12.2) | 69 (41-157) | 8.4 (5.4-13.9) | 68.6 (41.8-158.9) | <.001 |
| **Intensive BP Arm (%)** | 2824 (50.0%) | 606 (50.4%) | 619 (49.2%) | 203 (49.0%) | 0.91 |

Continuous variables are presented as mean (standard deviation) and categorical variables as count (percentage).
¶p-value by ANOVA for continuous variable or chi-square for categorical variables
\*UACR reported as median and 25-75<sup>th</sup> percentile.
ECG-LVG, electrocardiographic left ventricular hypertrophy; GFR, glomerular filtration rate; UACR, urine albumin creatinine ratio.

Over a median follow-up of 3.8 years (3.3-4.3), 182 cases of ADHF occurred. 8.7% of participants experienced ADHF events in (LVH + albuminuria) category compared to 1.2% in neither LVH, nor albuminuria category; 3.9% in isolated albuminuria, and 2.2% in the isolated LVH category. It is further depicted in Kaplan Meier curves, which showed the lowest ADHF survival in (LVH + albuminuria) category (Log-rank rank test p<0.001) **Figure 1**.

**Figure 1.**
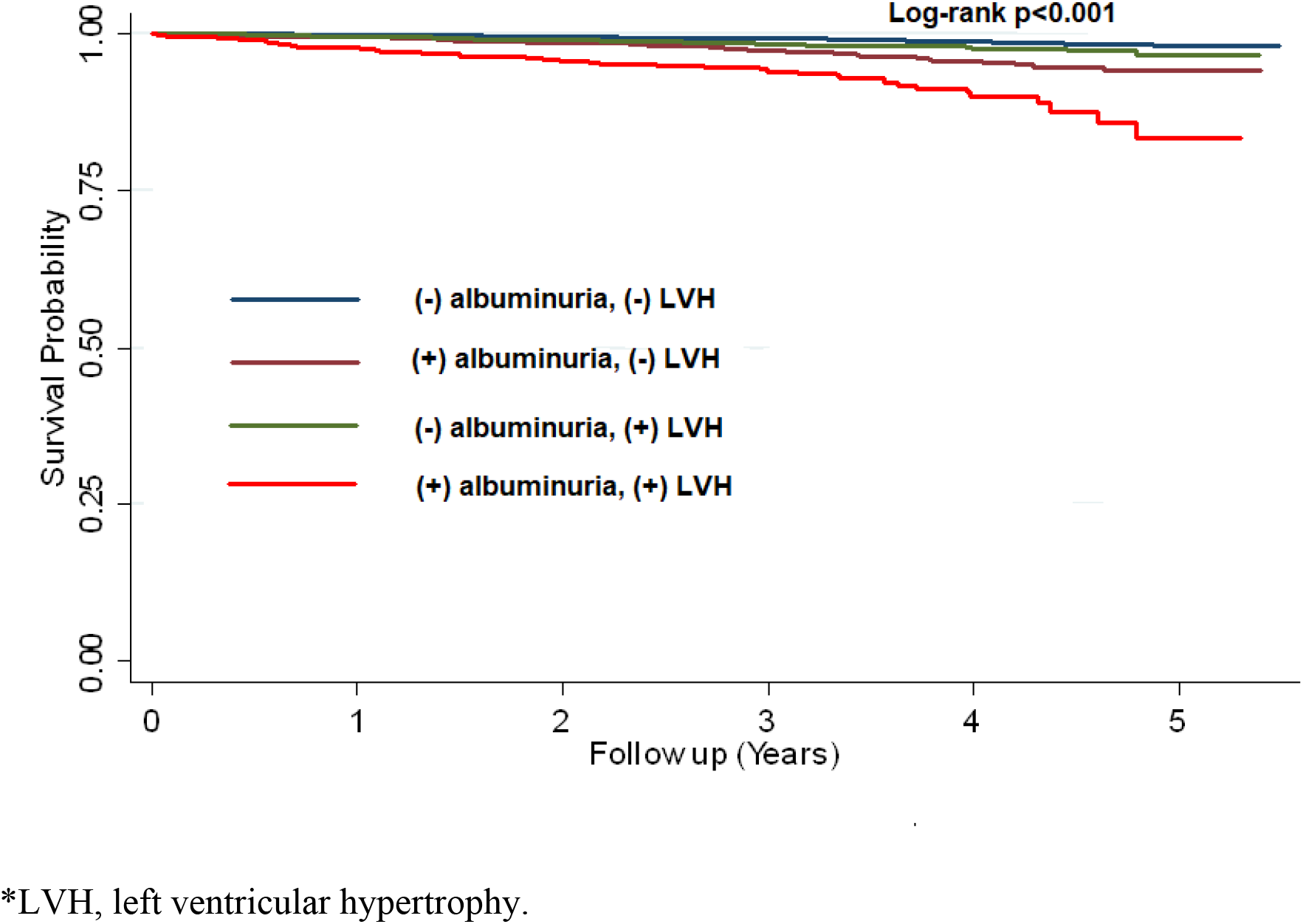
Kaplan Meier Curves of Incident ADHF by Albuminuria and Left Ventricular Hypertrophy Status*.

In multivariable-adjusted analysis, the presence of LVH vs. absence of LVH and albuminuria vs. no albuminuria were independently associated with ADHF (results not shown). There was a 5-fold higher risk of ADHF in the (+) albuminuria, (+) LVH category compared to (-) albuminuria, (-) LVH category (reference). Further, the (-) albuminuria, (+) LVH category was associated with a statistically non-significant 1.4-fold higher risk of ADHF, while (+) albuminuria, (-) LVH category was associated with a 2-fold higher risk of ADHF. (**Table 2**). There was significant additive interaction between albuminuria and LVH (relative excess risk due to interaction, 2.43; 95% confidence interval (CI): 0.53-4.33; *p-value* =0.01). However, there was no significant multiplicative interaction between albuminuria and LVH (*p-value* for interaction=0.11).

**Table 2.** Association of Albuminuria and LVH Categories with Incident ADHF.

| Albuminuria and LVH Categories | Events/Total | Model 1! | p-value | Model 2] | p-value |
| --- | --- | --- | --- | --- | --- |
|  | n (%) | HR (95% CI) |  | HR (95%CI) |  |
| **(-) Albuminuria (-) LVH** | 70/5638 (1.2%) | *Ref* | - | *Ref* | - |
| **(+) Albuminuria (-) LVH** | 48/1202 (3.9%) | 2.50 (1.72-3.63) | <.001 | 2.04 (1.39-3.00) | <.001 |
| **(-) Albuminuria (+) LVH** | 28/1257 (2.2%) | 1.78 (1.14-2.78) | 0.01 | 1.47 (0.93-2.32) | 0.09 |
| **(+) Albuminuria (+) LVH** | 36/414 (8.7%) | 6.80 (4.51-10.2) | <.001 | 4.95 (3.22-7.62) | <.001 |

! Model 1 adjusted for age, sex, race, and randomization site.
]Model 2 adjusted for smoking, body mass index, systolic blood pressure, total cholesterol, prevalent clinical/subclinical CVD, statin use, number of antihypertensive agents, eGFR, treatment assignment.
**Additive interaction p-value= 0.01.** Relative excess risk due to interaction, 2.43; 95% confidence interval (CI): 0.53-4.33; p=0.01).
Proportion of disease attributable to interaction (AP)= 0.49 (95% CI: 0.23-0.74)
Synergy index (S)= 2.60 (95% CI: 1.26-5.33)

The effect of intensive blood pressure control on ADHF risk appeared to be attenuated in (+) albuminuria, and (+) LVH category, however, albuminuria/LVH categories did not significantly modify the effects of intensive vs. standard blood pressure control on ADHF risk (*p-value* for interaction=0.26) (**Table 3**).

**Table 3.**
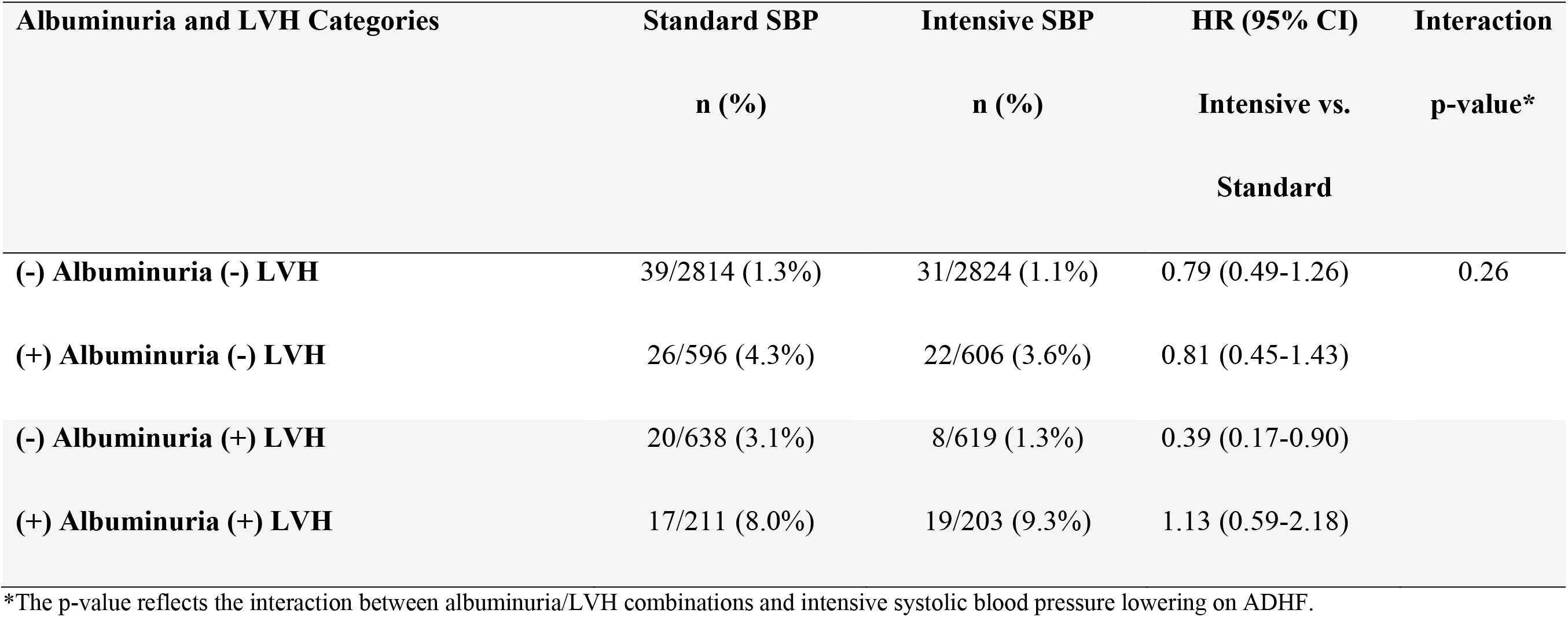
Effect of Intensive Blood Pressure Lowering on ADHF Stratified by Albuminuria and LVH combinations.

When albuminuria was defined as UACR ≥10 mg/g, similar results of a stronger association of the concomitant presence of albuminuria and LVH with incident ADHF than albuminuria and LVH in isolation was observed. (Additive and multiplicative interaction *p-value* between albuminuria (≥10 mg/g) and LVH = 0.01 and 0.048 respectively. (**Supplemental Table 1)**. The incidence rate of ADHF increased linearly across these UACR categories with a higher incidence rate in the presence of LVH **Figure 2**.

**Figure 2.**
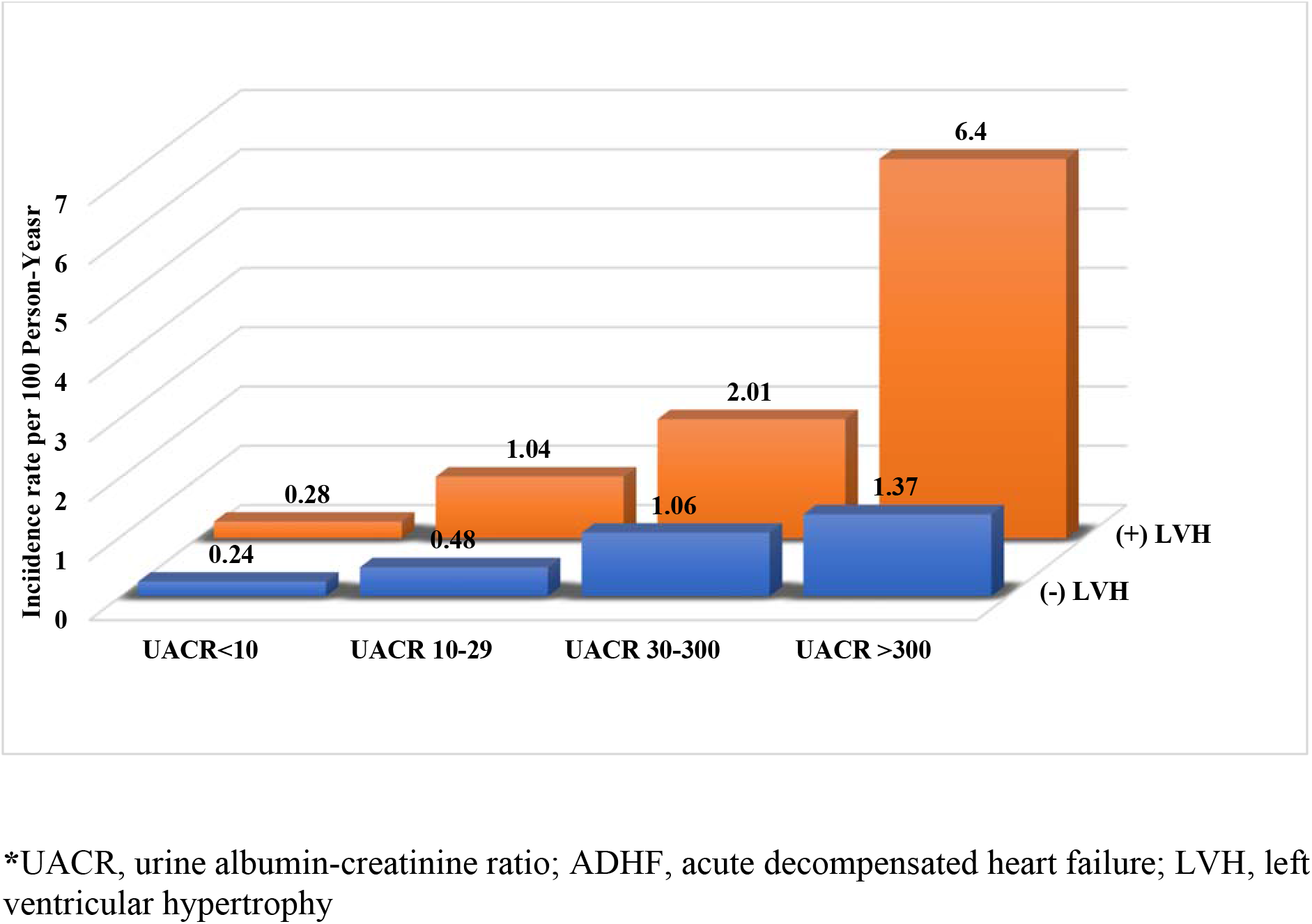
Incidence Rate of ADHF across UACR Categories Stratified by Left Ventricular Hypertrophy*.

In the multivariable-adjusted model compared to the reference group (UACR<10 mg/g and no LVH), the association between increasing UACR categories and incident ADHF was stronger in the presence of LVH than in the absence of LVH **(Table 4)**.

**Table 4.** Risk of ADHF by UACR Levels and LVH Groups.

| UACR and LVH Categories | Event/Total<br>n (%) | Model 1!<br>HR (95% CI) | p-value | Model 2]<br>HR (95%CI) | p-value |
| --- | --- | --- | --- | --- | --- |
| UACR <10 mg/g and (-) LVH | 35/3723 (0.94%) | *Ref* | - | *Ref* | - |
| UACR <10 mg/g and (+) LVH | 35/1915 (1.8%) | 1.64 (1.02-2.64) | 0.03 | 1.53 (0.95-2.46) | 0.07 |
| UACR 10-29.9 mg/g and (-) LVH | 8/739 (1.0%) | 1.14 (0.52-2.47) | 0.73 | 0.96 (0.44-2.08) | 0.91 |
| UACR 10-29.9 mg/g and (+) LVH | 20/518 (3.8%) | 3.63 (2.07-6.36) | <0.01 | 2.90 (1.63-5.16) | 0.001 |
| UACR 30-299.9 mg/g and (-) LVH | 40/1037 (3.8%) | 2.99 (1.89-4.75) | <.001 | 2.55 (1.59-4.09) | <.001 |
| UACR 30-299.9 mg/g and (+) LVH | 25/365 (7.0%) | 6.65 (3.95-11.20) | <.001 | 4.92 (2.87-8.43) | <.001 |
| UACR ≥300 mg/g and (-) LVH | 8/165 (4.8%) | 4.09 (1.89-8.85) | 0.01 | 2.58 (1.12-5.95) | 0.02 |
| UACR ≥300 mg/g and (+) LVH | 11/58 (18.9%) | 24.8 (12.4-49.3) | <.001 | 17.9 (8.59-37.3) | <.001 |

! Model 1 adjusted for age, sex, race, and randomization site.
]Model 2 adjusted for smoking, body mass index, systolic blood pressure, total cholesterol, prevalent clinical/subclinical CVD, statin use, number of antihypertensive agents, eGFR, and treatment assignment.
UACR; urine albumin-creatinine ratio; LVH; left ventricular hypertrophy.

**Supplemental Tables 2, 3, and 4** show multivariable-adjusted models of the association of LVH/albuminuria combinations with ADHF using Cornell Voltage, Sokolow Lyon, and Cornell product criteria respectively. Similar, higher incidence and relative risk of ADHF was observed with (+) LVH, (+) albuminuria combination compared to (-) LVH, (-) albuminuria (reference).

## Discussion

In this post-hoc analysis from the SPRINT trial, we evaluated the independent and joint association of LVH and albuminuria with incident HF. The main finding of our analysis from this trial was that LVH and albuminuria were independent and additive predictors of HF risk with significant interaction between these predictors. The combination of LVH and albuminuria posed the highest risk of ADHF than LVH or albuminuria in isolation and the risk increased with increasing levels of albuminuria. Additionally, the effect of intensive blood pressure control in decreasing ADHF risk did not vary significantly across albuminuria/LVH combinations. Taken together, these findings suggest that LVH and albuminuria even at low-grade levels are strong additive predictors of future ADHF and therefore, the concomitant presence of these markers may identify high-risk individuals in need of intensive surveillance and neurohormonal modification with potential newer therapies.

Albuminuria is prevalent in high-risk populations such as those with obesity, diabetes mellitus, and hypertension and is increasingly being recognized as an important predictor of HF^6,17^. Albuminuria reflects structural damage to the filtration barrier and is linked to CVD through multifactorial mechanisms such as endothelial damage, systemic inflammation, presence of comorbid conditions, and neurohormonal activation, including activation of the renin-angiotensin-aldosterone system (RAAS)^18^. Systemic inflammation and neurohormonal activation via the heightened activity of RAAS promote oxidative stress and more systemic vascular injury. These pathophysiological processes may promote volume overload via an increase in sodium and water retention and cardiac filling pressures due to increased reabsorption, thereby promoting the development and progression of HF^6^. The resulting venous congestion may promote renal venous congestion, resulting in a decline in effective perfusion pressure and eGFR which may lead to further activation of compensatory mechanisms such as RASS and thus creating a bidirectional progressive cardiorenal injury^19^. Therefore, albuminuria is not only a predictor of future HF in otherwise healthy participants but among patients with HF, albuminuria is an independent predictor of exacerbation of HF and increased mortality risk^20-22^. Similarly, LVH is an established risk for HF and despite the low sensitivity of ECG to detect LV mass, ECG-LVH is still a strong predictor of HF^23^. Individuals with asymptomatic LVH who were later transitioned to HF are hypothesized to have maladaptive cardiac remodeling due to myocardial injury, inflammation, and fibrosis^23,24^ and albuminuria is one of the pathophysiological links between LVH and inflammation^25^. On the other hand, albuminuria is associated with eccentric and concentric LVH, and this abnormal LV geometry may mediate the association of albuminuria with CVD events^26,27^. Analysis of data from the Losartan Intervention For Endpoint reduction in hypertension (LIFE) study showed that both LVH and albuminuria independently increased the risk of CVD events and mortality.^28^ Additionally, the risk of CVD outcomes with albuminuria appeared to occur at much lower levels of UACR among participants with hypertension and LVH.^29^ Thus, both albuminuria and LVH which represent long-term consequences of uncontrolled hypertension are strongly interrelated in a complex and bidirectional relationship and may modify/amplify the effect of each other in predicting CVD outcomes as observed in our study.

In this study, we did not find evidence that baseline albuminuria and LVH modified the effects of intensive systolic BP lowering on HF incidence. The effect of intensive systolic blood pressure lowering on the reduction of HF incidence was attenuated among participants with concomitant LVH and albuminuria. Notably, a prior post-hoc analysis from SPRINT trial also reported that albuminuria did not modify the absolute benefits and risks of intensive systolic BP lowering.^30^Long-term follow-up data is needed to draw further conclusions on the long-term effect of intensive systolic BP lowering on the risk of HF among participants with concomitant LVH and albuminuria.

Albuminuria and LVH are both potentially modifiable risk factors and regression of these markers has been associated with a reduction of subsequent CVD events^10,31^. For example, in addition to RAAS blockade, sodium-glucose cotransporter 2 inhibitors (SGLT2i) and nonsteroidal mineralocorticoid receptor antagonists (MRAs) have been proven effective in reducing the progression of albuminuria, with resultant reduction in the progression of chronic kidney disease (CKD) and HF hospitalization ^8,32,33^. Similarly, regression of LVH by ECG or imaging with antihypertensive medications has been associated with a reduced risk of CVD events including HF^10,34^. Although universal screening for LVH is not recommended due to the paucity of data on the value of ECG in assessing CVD risk^35^, our findings suggest that wider availability of ECG may render it as an effective tool that can be employed for surveillance in high-risk populations such as those with albuminuria to identify participants in need of aggressive risk factor control. On the other hand, screening for albuminuria also represents a scalable and cost-effective method to detect and prevent CVD but despite a promising marker for risk stratification, testing for spot albuminuria remains suboptimal in routine clinical care^36^. Therefore, regular screening of albuminuria especially in participants with diabetes mellitus and hypertension may prevent the progression of cardiorenal disease with early interventions ^37^.

### Study Limitations and Strengths

Certain limitations of the study are the following: First, we only used baseline measurement of LVH and albuminuria and did not take into account their progression or regression during the follow-up period. Secondly, we lacked imaging data for LVH in SPRINT, accordingly ECG criteria which may be a less sensitive method than echocardiography or cardiac magnetic resonance^5^. However, LVH detected by ECG has similar prognostic value in predicting poor outcomes as LVH detected by imaging^38^. Thirdly, overall fewer HF events and the exclusion of new outpatient HF as an endpoint event may limit the statistical power of the analysis. Fourthly, our results may not be generalizable to other populations who did not meet eligibility criteria for SPRINT such as individuals with diabetes mellitus, individuals with low CVD risk, prior stroke, <50 years old, and individuals residing in nursing homes or assisted living facilities. The strengths of our study include a large sample size from the well-designed clinical trial of a diverse population at high risk of CVD. The data variables were collected using standardized procedures in the context of a clinical trial with the central reading of ECGs blinded to the treatment assignment. Since ADHF was a component of the primary composite outcome, robust protocol-driven procedures were employed to ensure the validity of ADHF events in SPRINT.

## Conclusion

In SPRINT, albuminuria and LVH are independent and additive predictors of ADHF, and the incidence and relative risk of ADHF increases with increasing levels of albuminuria, with the highest risk in the presence of LVH than in the absence of it. The concomitant presence of LVH and albuminuria can potentially identify participants who need intensive disease management to prevent HF.

### Clinical Perspective

Albuminuria and LVH are independent and additive predictors of ADHF and the relative risk of ADHF is progressively higher with increasing levels of albuminuria with a higher risk in the presence of LVH than in the absence of it. Albuminuria and LVH are both potentially modifiable risk factors as regression of these markers have been associated with reduced risk of future CVD including HF. Whether intensive blood pressure control and modification of neurohormonal activation with newly approved therapies would reduce the risk of ADHF among participants with concomitant albuminuria and LVH need to be tested in large clinical trials with longer-follow up.

### Novelty And Relevance

What is New?

This is the first study to directly examine the interaction between albuminuria and LVH on ADHF outcome in participants with hypertension.

What is Relevant?

Albuminuria and LVH are additive predictors of ADHF, and the risk increases with increasing levels of albuminuria especially in those with concomitant LVH.

Clinical/Pathophysiological Implication?

Both albuminuria and LVH are potentially modifiable risk factors and regression of these markers have been associated with reduced risk of CVD outcomes including HF. Individuals with concomitant albuminuria and LVH may need intensive surveillance and consideration of newer disease modifying therapies.

## Data Availability

All data and materials have been made publicly available at the National Heart, Lung, and Blood Institute BioLINCC data repository and can be accessed at https://biolincc.nhlbi.nih.gov/home/.

## Funding

None.

## Disclosures

Dr. Chen receives funding from the National Heart, Lung, and Blood Institute (NHLBI) (R01 HL141288, R01 HL126637, K24 HL155813). The remaining authors have no disclosures to report.

## Notes

### Competing Interest Statement

The authors have declared no competing interest.

### Author Declarations

Medical College of Wisconsin IRB

## References

1. Staplin N, Haynes R, Herrington WG. Blood pressure and kidney disease: chicken or egg (or both)? Kidney International. 2020/09/01/ 2020;98(3):547-549. doi:https://doi.org/10.1016/j.kint.2020.05.048

2. Viazzi F, Pontremoli R. Blood pressure, albuminuria and renal dysfunction: the ‘chicken or egg’ dilemma. Nephrol Dial Transplant. Aug 2014;29(8):1453–5. doi:10.1093/ndt/gfu183

3. Kramer H, Jacobs DR, Jr., Bild D, et al. Urine albumin excretion and subclinical cardiovascular disease. The Multi-Ethnic Study of Atherosclerosis. Hypertension. Jul 2005;46(1):38–43. doi:10.1161/01.Hyp.0000171189.48911.18

4. Palmieri V, Tracy RP, Roman MJ, et al. Relation of left ventricular hypertrophy to inflammation and albuminuria in adults with type 2 diabetes: the strong heart study. Diabetes Care. Oct 2003;26(10):2764–9. doi:10.2337/diacare.26.10.2764

5. Oseni AO, Qureshi WT, Almahmoud MF, et al. Left ventricular hypertrophy by ECG versus cardiac MRI as a predictor for heart failure. Heart. 2017;103(1):49–54. doi:10.1136/heartjnl-2016-309516

6. Khan MS, Shahid I, Anker SD, et al. Albuminuria and Heart Failure: JACC State-of-the-Art Review. Journal of the American College of Cardiology. 2023/01/24/ 2023;81(3):270-282. doi:https://doi.org/10.1016/j.jacc.2022.10.028

7. Bailey LN, Levitan EB, Judd SE, et al. Association of Urine Albumin Excretion With Incident Heart Failure Hospitalization in Community-Dwelling Adults. JACC Heart Fail. May 2019;7(5):394–401. doi:10.1016/j.jchf.2019.01.016

8. August P. Chronic Kidney Disease — Another Step Forward. New England Journal of Medicine. 2023;388(2):179–180. doi:10.1056/NEJMe2215286

9. Soliman EZ, Ambrosius WT, Cushman WC, et al. Effect of Intensive Blood Pressure Lowering on Left Ventricular Hypertrophy in Patients With Hypertension: SPRINT (Systolic Blood Pressure Intervention Trial). Circulation. Aug 1 2017;136(5):440–450. doi:10.1161/circulationaha.117.028441

10. Mathew J, Sleight P, Lonn E, et al. Reduction of cardiovascular risk by regression of electrocardiographic markers of left ventricular hypertrophy by the angiotensin-converting enzyme inhibitor ramipril. Circulation. Oct 2 2001;104(14):1615–21. doi:10.1161/hc3901.096700

11. Ambrosius WT, Sink KM, Foy CG, et al. The design and rationale of a multicenter clinical trial comparing two strategies for control of systolic blood pressure: the Systolic Blood Pressure Intervention Trial (SPRINT). Clin Trials. Oct 2014;11(5):532–46. doi:10.1177/1740774514537404

12. Levey AS, Bosch JP, Lewis JB, Greene T, Rogers N, Roth D. A more accurate method to estimate glomerular filtration rate from serum creatinine: a new prediction equation. Modification of Diet in Renal Disease Study Group. Ann Intern Med. Mar 16 1999;130(6):461–70. doi:10.7326/0003-4819-130-6-199903160-00002

13. Molloy TJ, Okin PM, Devereux RB, Kligfield P. Electrocardiographic detection of left ventricular hypertrophy by the simple QRS voltage-duration product. Journal of the American College of Cardiology. 1992/11/01/ 1992;20(5):1180-1186. doi:https://doi.org/10.1016/0735-1097(92)90376-X

14. Chang AR, Kramer H, Wei G, et al. Effects of Intensive Blood Pressure Control in Patients with and without Albuminuria: Post Hoc Analyses from SPRINT. Clin J Am Soc Nephrol. Aug 7 2020;15(8):1121–1128. doi:10.2215/cjn.12371019

15. Rosamond WD, Chang PP, Baggett C, et al. Classification of heart failure in the atherosclerosis risk in communities (ARIC) study: a comparison of diagnostic criteria. Circ Heart Fail. Mar 1 2012;5(2):152–9. doi:10.1161/circheartfailure.111.963199

16. Li R, Chambless L. Test for Additive Interaction in Proportional Hazards Models. Annals of Epidemiology. 2007/03/01/ 2007;17(3):227-236. doi:https://doi.org/10.1016/j.annepidem.2006.10.009

17. Garg AX, Kiberd BA, Clark WF, Haynes RB, Clase CM. Albuminuria and renal insufficiency prevalence guides population screening: Results from the NHANES III. Kidney International. 2002/06/01/ 2002;61(6):2165-2175. doi:https://doi.org/10.1046/j.1523-1755.2002.00356.x

18. Weir MR. Microalbuminuria and cardiovascular disease. Clin J Am Soc Nephrol. May 2007;2(3):581–90. doi:10.2215/cjn.03190906

19. Damman K, Deursen VMv, Navis G, Voors AA, Veldhuisen DJv, Hillege HL. Increased Central Venous Pressure Is Associated With Impaired Renal Function and Mortality in a Broad Spectrum of Patients With Cardiovascular Disease. Journal of the American College of Cardiology. 2009;53(7):582–588. doi:doi:10.1016/j.jacc.2008.08.080

20. Damman K, Valente MA, Voors AA, O’Connor CM, van Veldhuisen DJ, Hillege HL. Renal impairment, worsening renal function, and outcome in patients with heart failure: an updated meta-analysis. Eur Heart J. Feb 2014;35(7):455–69. doi:10.1093/eurheartj/eht386

21. Dries DL, Exner DV, Domanski MJ, Greenberg B, Stevenson LW. The prognostic implications of renal insufficiency in asymptomatic and symptomatic patients with left ventricular systolic dysfunction. Journal of the American College of Cardiology. 2000;35(3):681–689. doi:doi:10.1016/S0735-1097(99)00608-7

22. Selvaraj S, Claggett B, Shah SJ, et al. Prognostic Value of Albuminuria and Influence of Spironolactone in Heart Failure With Preserved Ejection Fraction. Circ Heart Fail. Nov 2018;11(11):e005288. doi:10.1161/circheartfailure.118.005288

23. Almahmoud MF, O’Neal WT, Qureshi W, Soliman EZ. Electrocardiographic Versus Echocardiographic Left Ventricular Hypertrophy in Prediction of Congestive Heart Failure in the Elderly. Clin Cardiol. Jun 2015;38(6):365–70. doi:10.1002/clc.22402

24. Kannel WB, D’Agostino RB, Silbershatz H, Belanger AJ, Wilson PW, Levy D. Profile for estimating risk of heart failure. Arch Intern Med. Jun 14 1999;159(11):1197–204. doi:10.1001/archinte.159.11.1197

25. Palmieri V, Tracy RP, Roman MJ, et al. Relation of Left Ventricular Hypertrophy to Inflammation and Albuminuria in Adults With Type 2 Diabetes: The Strong Heart Study. Diabetes Care. 2003;26(10):2764–2769. doi:10.2337/diacare.26.10.2764

26. Koren MJ, Devereux RB, Casale PN, Savage DD, Laragh JH. Relation of left ventricular mass and geometry to morbidity and mortality in uncomplicated essential hypertension. Ann Intern Med. Mar 1 1991;114(5):345–52. doi:10.7326/0003-4819-114-5-345

27. Wachtell K, Palmieri V, Olsen MH, et al. Urine albumin/creatinine ratio and echocardiographic left ventricular structure and function in hypertensive patients with electrocardiographic left ventricular hypertrophy: the LIFE study. Losartan Intervention for Endpoint Reduction. Am Heart J. Feb 2002;143(2):319–26. doi:10.1067/mhj.2002.119895

28. Wachtell K, Ibsen H, Olsen MH, et al. Albuminuria and cardiovascular risk in hypertensive patients with left ventricular hypertrophy: the LIFE study. Ann Intern Med. 2003;139(11):901–906.

29. Olsen MH, Wachtell K, Ibsen H, et al. Reductions in albuminuria and in electrocardiographic left ventricular hypertrophy independently improve prognosis in hypertension: the LIFE study. J Hypertens. 2006;24(4):775–781.

30. Chang AR, Kramer H, Wei G, et al. Effects of intensive blood pressure control in patients with and without albuminuria: post hoc analyses from SPRINT. Clin J Am Soc Nephrol. 2020;15(8):1121–1128.

31. Herrington WG, Staplin N, Wanner C, et al. Empagliflozin in Patients with Chronic Kidney Disease. N Engl J Med. Jan 12 2023;388(2):117–127. doi:10.1056/NEJMoa2204233

32. Bakris GL, Ruilope LM, Anker SD, et al. A prespecified exploratory analysis from FIDELITY examined finerenone use and kidney outcomes in patients with chronic kidney disease and type 2 diabetes. Kidney International. 2023/01/01/ 2023;103(1):196-206. doi:https://doi.org/10.1016/j.kint.2022.08.040

33. Navaneethan SD, Zoungas S, Caramori ML, et al. Diabetes Management in Chronic Kidney Disease: Synopsis of the KDIGO 2022 Clinical Practice Guideline Update. Ann Intern Med. Jan 10 2023;doi:10.7326/m22-2904

34. Okin PM, Devereux RB, Jern S, et al. Regression of Electrocardiographic Left Ventricular Hypertrophy During Antihypertensive Treatment and the Prediction of Major Cardiovascular Events. JAMA. 2004;292(19):2343–2349. doi:10.1001/jama.292.19.2343

35. Whelton PK, Carey RM, Aronow WS, et al. 2017 ACC/AHA/AAPA/ABC/ACPM/AGS/APhA/ASH/ASPC/NMA/PCNA Guideline for the Prevention, Detection, Evaluation, and Management of High Blood Pressure in Adults. Journal of the American College of Cardiology. 2018;71(19):e127-e248. doi:doi:10.1016/j.jacc.2017.11.006

36. Shin JI, Chang AR, Grams ME, et al. Albuminuria Testing in Hypertension and Diabetes: An Individual-Participant Data Meta-Analysis in a Global Consortium. Hypertension. Sep 2021;78(4):1042–1052. doi:10.1161/hypertensionaha.121.17323

37. Alfego D, Ennis J, Gillespie B, et al. Chronic Kidney Disease Testing Among At-Risk Adults in the U.S. Remains Low: Real-World Evidence From a National Laboratory Database. Diabetes Care. Sep 2021;44(9):2025–2032. doi:10.2337/dc21-0723

38. Leigh JA, O’Neal WT, Soliman EZ. Electrocardiographic Left Ventricular Hypertrophy as a Predictor of Cardiovascular Disease Independent of Left Ventricular Anatomy in Subjects Aged ≥65 Years. Am J Cardiol. Jun 1 2016;117(11):1831–5. doi:10.1016/j.amjcard.2016.03.020

